# Proportion of idiopathic pulmonary fibrosis risk explained by known genetic loci

**DOI:** 10.1101/2020.08.14.20172528

**Authors:** Olivia C Leavy, Shwu-Fan Ma, Philip L Molyneaux, Toby M Maher, Justin M Oldham, Carlos Flores, Imre Noth, R Gisli Jenkins, Frank Dudbridge, Louise V Wain, Richard J Allen

## Abstract

Genome-wide association studies have identified 14 genetic loci associated with susceptibility to idiopathic pulmonary fibrosis (IPF), a devastating lung disease with poor prognosis. Of these, the variant with the strongest association, rs35705950, is located in the promoter region of the *MUC5B* gene and has a risk allele (T) frequency of 30-35% in IPF cases. Here we present estimates of the proportion of disease liability explained by each of the 14 IPF risk variants as well as estimates of the proportion of cases that can be attributed to each variant. We estimate that rs35705950 explains 5.9-9.4% of disease liability, which is much lower than previously reported estimates. Of every 100,000 individuals with the rs35705950_GG genotype we estimate 30 will have IPF, whereas for every 100,000 individuals with the rs35705950_GT genotype 152 will have IPF. Quantifying the impact of genetic risk factors on disease liability improves our understanding of the underlying genetic architecture of IPF and provides insight into the impact of genetic factors in risk prediction modelling.

## Introduction

Understanding how genetic factors contribute to disease risk improves our understanding of pathogenesis, supports drug development and aids risk prediction^1^. Appropriate quantification and interpretation of this contribution is essential for measuring the impact of genetic variation and in motivating and informing future studies.

Idiopathic pulmonary fibrosis (IPF) is a chronic disease characterised by scarring of the lungs. Current therapies only slow disease progression and half of individuals die within 3-5 years of diagnosis. A genetic variant, rs35705950, in the *MUC5B* promoter region, is strongly associated with IPF susceptibility with the risk allele (T) associated with a five-fold increase in disease risk^2^. Genome-wide association studies (GWAS) have identified 13 additional independent IPF susceptibility variants^3^.

The rs35705950_T allele frequency in IPF cases is 30-35%^4^ (compared to 11% in controls), but risk allele frequency does not reflect the disease risk accounted for by this variant. Explained risk can be measured in different ways, such as the proportion of risk explained in the general population, or alternatively the proportion of cases due to a specific variant.

Here we provide estimates of the proportion of IPF risk in the general population explained by known IPF susceptibility variants, and estimates of the proportion of cases attributable to each susceptibility variant. Our analyses focussed on non-familial IPF, therefore variants considered are just those evidenced by GWAS.

## Methods

We used unrelated European IPF cases (diagnosed according to international guidelines^5^) and controls from a previously reported study^3^ with appropriate ethics approval.

To estimate the proportion of disease risk explained by each variant in the general population, we performed regression analyses including the susceptibility variant as the only covariate. R^2^ is a measure of phenotypic variance explained by a model and, as our model only contains a single variant, the proportion of disease explained by that variant. R^2^ cannot be directly calculated as the IPF phenotype is binary and the proportion of cases in our analysis is higher than observed in the general population. We therefore calculated a liability R^2^ accounting for enrichment of cases^6^. The liability model assumes individuals have an unmeasured continuous trait, called the liability, and an individual develops IPF when the liability exceeds a critical value. We calculated the liability R^2^ for IPF prevalence estimates of 1.25 and 63 cases per 100,000 people (the lowest and highest reported estimates of disease prevalence^7^). To estimate the variance in the liability explained by all variants, we fitted the model with the most significant variants from all 14 known IPF susceptibility loci and calculated the liability R^2^. Finally, we fitted the model with all susceptibility variants, minus rs35705950. We investigated whether results were biased by population stratification by repeating analyses including 10 genetic principal components to adjust for ancestry.

To estimate the proportion of cases attributable to each variant, we calculated the population attributable risk fraction^8^ (PARF). PARF is the proportion of cases that would be prevented if a risk factor were removed from the population. If any risk factors were removed, the PARFs of other risk factors would change. Therefore, PARFs cannot be summed to calculate the proportion of cases prevented if multiple risk factors were removed.

## Results

A total of 792 IPF cases and 10,000 controls were included in the analysis. Variant rs35705950 alone explains 5.9-9.4% of disease liability in the general population. No other IPF susceptibility variant explained more than 0.6% and collectively the 13 *non-MUC5B* susceptibility variants explained 1.8-2.9% of variation in disease liability (**Figure 1**). The highest PARF was observed for rs35705950 (51%), however many of the susceptibility variants had PARF>10 (**Figure 2**). Effect sizes were similar after adjusting for principal components suggesting results are not biased by population stratification.

**Figure 1:**
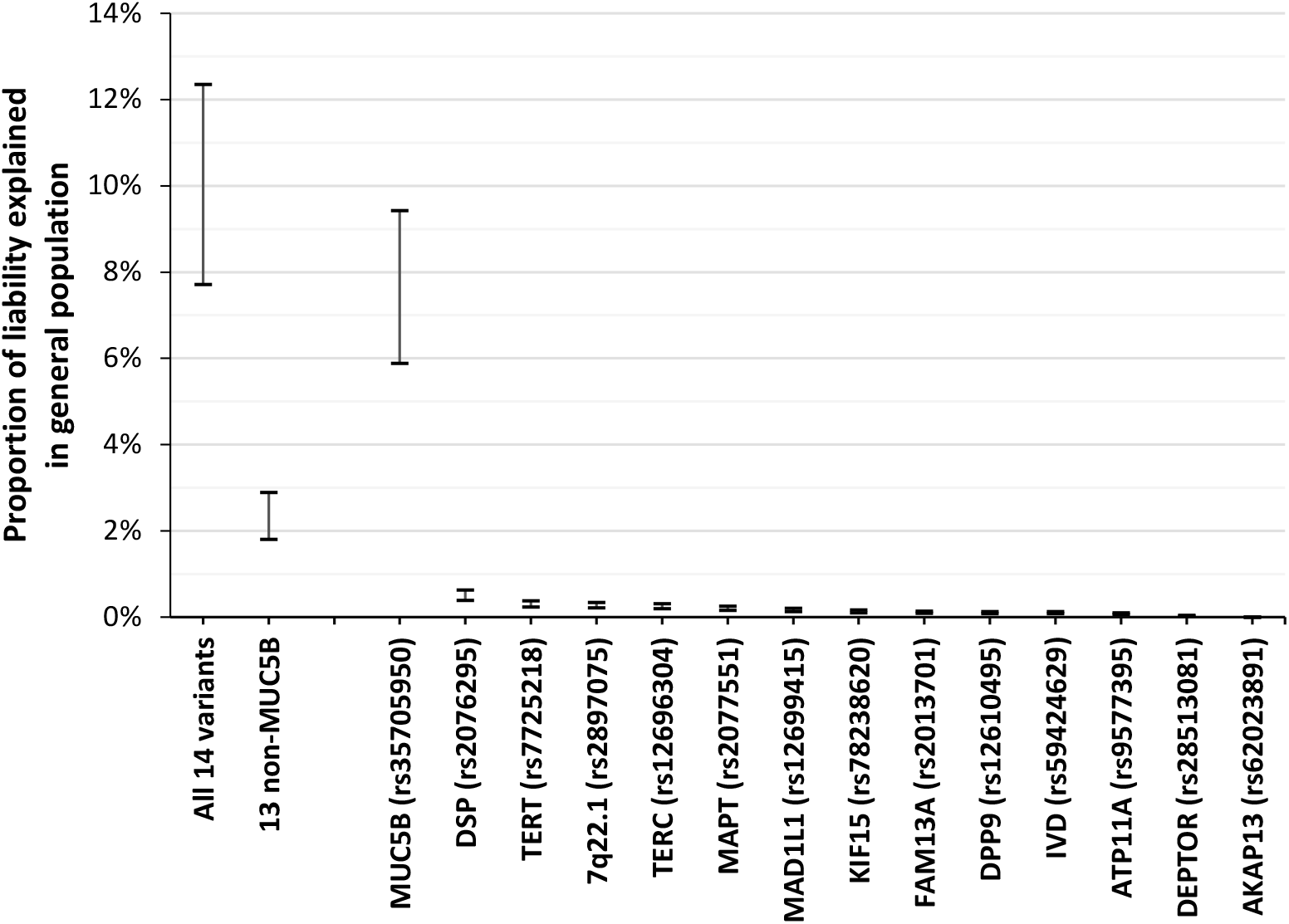
Proportion of liability explained in the general population. Estimated proportion of variation explained is taken from the liability R^2^ from the regression analyses with the lower bound given when assuming a disease prevalence of 1.25 cases per 100,000 and the upper bound given when assuming a disease prevalence of 63 cases per 100,000. The x-axis label “All 14 variants” refers to the model including all 14 sentinel IPF susceptibility variants, “13 non-MUC5B” refers to the model including all sentinel IPF susceptibility variants minus the *MUC5B* polymorphism rs35705950. Variants are ordered by the proportion of explained variation.

**Figure 2:**
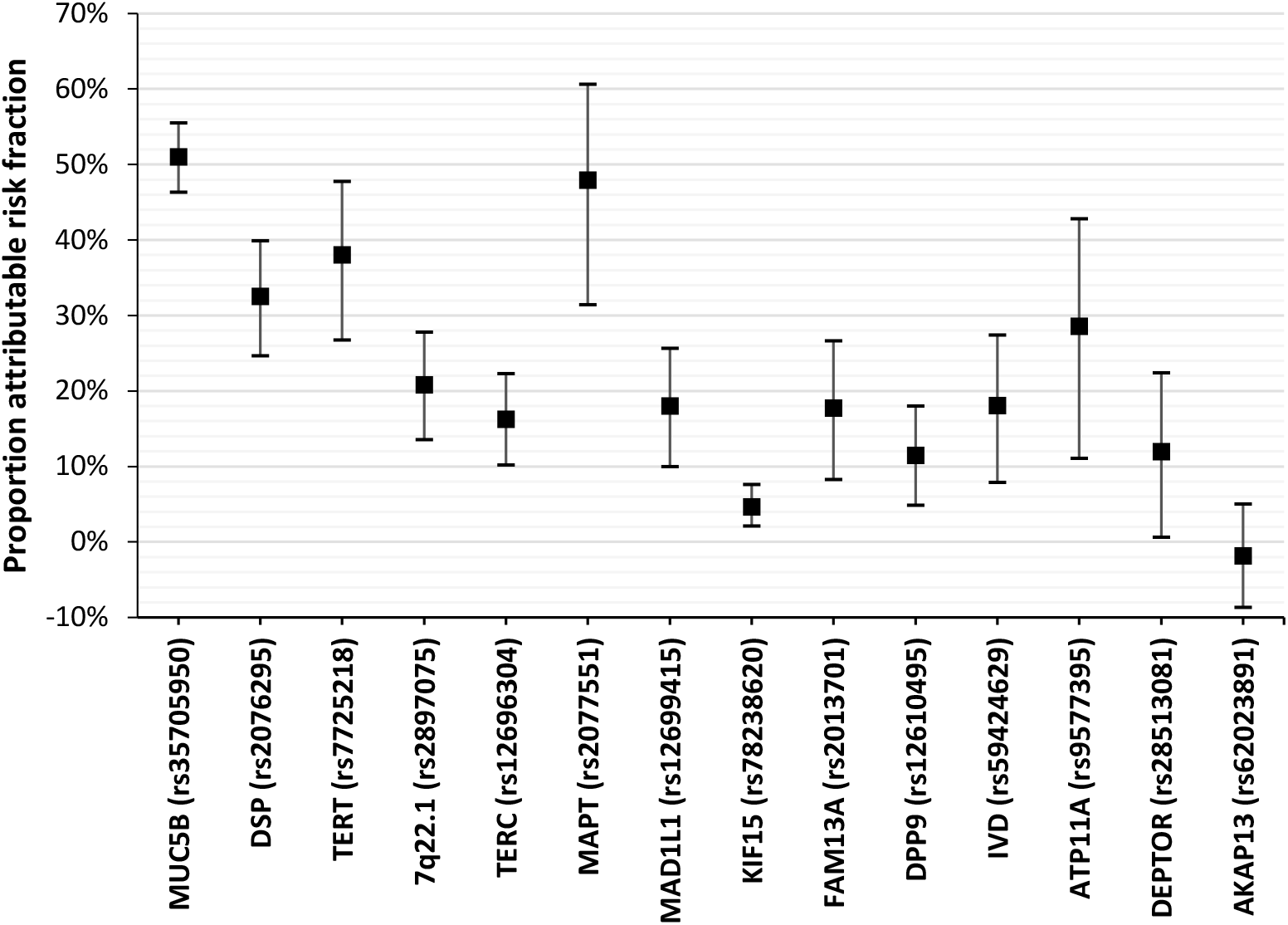
Population attributable risk fraction. Estimates of PARF for each variant with 95% confidence intervals. Variants are ordered by the proportion of explained variation in the general population (Figure 1).

## Discussion

The *MUC5B* promoter polymorphism explains three times more disease liability in the general population than the other 13 IPF susceptibility variants combined. In total, the 14 IPF susceptibility variants explain up to 12.4% of disease liability in the general population, which is smaller than previous reports that cited 30-35% of risk^4,9^. Importantly, therapies that target variants that explain a small proportion of disease risk can still have a large clinical impact^1^.

Our results suggest IPF cases could be halved if the *MUC5B* risk allele was removed from the population. Although the clinical relevance of PARF estimates may be limited as removing risk alleles from the general population is almost impossible, they do indicate the impact preventative interventions could have on disease incidence.

Different populations experience diverse environmental exposures and have varying allele frequencies, affecting the proportion of risk explained by these variants. We also only investigated known common IPF susceptibility variants, though previous studies suggest there could be many undiscovered genetic variants contributing to IPF risk^3^, and we have not investigated epistasis or gene-environment interactions. This means overall IPF risk explained by genetics will likely be much higher than the 12.4% explained by the known variants.

This study utilised an ascertained case-control study design and made assumptions about disease prevalence. Ideally, a general population cohort, such as UK Biobank^10^, would be used for these analyses. However, in UK Biobank there are few self-reported cases (n=104) and cases defined using hospital episode statistics do not genetically resemble clinically recruited cases (rs35705950_T allele frequency in these cases is 20%). Therefore, we restricted analyses to a study with clinically recruited cases. The study used was not used in the discovery of the IPF susceptibility variants^3^, meaning the estimates of risk explained should not be subject to winner’s curse bias.

We could also consider absolute risk. Assuming disease prevalence is 63 cases per 100,000 and using the previously reported^2^ effect size for the *MUC5B* risk allele (OR=4.99), for every 100,000 individuals with the rs35705950_GG genotype we would expect 30 to have IPF, whereas for every 100,000 individuals with the rs35705950_GT genotype we would expect 152 to have IPF. Therefore, although rs35705950 is strongly associated with disease risk, most individuals carrying the risk allele will not develop IPF.

Although risk allele frequencies in cases can be of interest, they are not a measure of explained risk. Many of the known IPF susceptibility variants have a high PARF, but individually explain a small overall proportion of the variation in risk. These results provide an important reference point to inform future genetic discoveries and for evaluation of the likely contribution of genetic factors in risk prediction models.

## Data Availability

All results are presented in the paper in Figures 1 and 2. Further enquires about data can be made to the corresponding author.

